# Systematic review of instruments for assessing culinary skills in adults: What is the quality of their psychometric properties?

**DOI:** 10.1101/2020.06.12.20129668

**Authors:** Aline Rissatto Teixeira, Daniela Bicalho, Betzabeth Slater, de Mendonça Lima Tacio

## Abstract

**Background:** Culinary skills are important objects of study in the field of Public Health. Studies that propose to develop instruments for assessing such construct show lack of methodological uniformity to report validity and reliability of their instruments.

**Objective:** To identify studies that have developed instruments to measure culinary skills in adult population, and critically assess their psychometric properties.

**Design:** We conducted a systematic review according to the PRISMA statement. We searched literature PubMed/Medline, Scopus, LILACS, and Web of Science databases until January 2021, and consulted Google Scholar for relevant grey literature. Two reviewers independently selected the studies, conducted data extraction, and assessed the psychometric quality of the instruments. A third reviewer resolved any doubts or disagreements in all steps of the systematic review.

**Results:** The search identified 1148 potentially relevant studies, out of which 9 met the inclusion criteria. In addition, we included 3 studies by searching the related articles and the reference lists of these studies, totaling 12 included studies in this review. Ten studies reported the development of tools measuring culinary skills in adults and 2 studies performed cross-cultural adaptations of original instruments. We considered adequate quality of internal consistency reliability in four studies. One study received adequate rating for test-retest reliability. No studies presented adequate rating for content validity and four studies showed satisfactory results for at least one type of construct validity. One study reported criterion validity and the quality of this psychometric property was inadequate.

**Conclusions:** We identified many studies that surveyed culinary skills. Although the isolated measures appraised in this review show good promise in terms of quality of psychometric properties, no studies presented adequate measures for each aspect of reliability and validity. A more consistent and consensual definition of culinary skills is recommended. The flaws observed in these studies show that there is a need for ongoing research in the area of the psychometric properties of instruments assessing culinary skills.

## Introduction

The discussion about the improvement of culinary skills and food practices has proven to be an important object of study in the field of Public Health; these skills are key factors associated with eating behaviors and with several complexities that represent social determinants of health [1].

Several authors define the term culinary skills in their publications [2-6], however, there is no consensus on its definition or a consistent theoretical debate about it [3]. This systematic review considers a broad definition of culinary skills proposed by De Oliveira, 2018 [7], as a set of attributes related to the selection and combination of foods and the use of culinary procedures and utensils involved in the planning, organization and preparation of “from scratch” meals based on fresh, minimally processed foods and culinary ingredients.

Culinary skills are associated with other concepts that involve the practice of proper and healthy eating, such as food literacy, which takes into account the broader social and environmental dimensions of eating together, associated with an individual’s abilities [8]. Those considered to be “food literate” have the skills and abilities to revise and adapt their diet and food sources in response to changes imposed by modern life to maintain dietary quality [8]. Another concept related to culinary skills is food agency, which relates to the ability to act intentionally to change their own food environment. In general, its focus is on the individual mechanisms that lead to the act of cooking at home, secondary to other external elements that impact on the freedom of the individual and, consequently, on their autonomy [9]. Culinary autonomy is defined as the ability to think, decide, and act, to cook meals at home using mostly fresh and minimally processed foods, under the influence of interpersonal relationships, the environment, cultural values, access to opportunities, and the guarantee of rights; therefore, culinary skills represent an important dimension of this construct [7].

Time devoted to cooking has decreased and has been viewed as a global trend: food industry investments in advertising and marketing to “solve the everyday food problem” devalue cooking as an emancipatory competence associated with a healthy food routine [10]. Such decrease is associated with greater purchase of ultra-processed foods, and concerns public health experts around the world, considering their negative nutritional attributes and harmful effects on consumers’ health, such as overweight, obesity, cancer and other chronic diseases and addiction-like behavior [11; 12]. It is worth mentioning that culinary practices also relate to environmental, social and economic implications. Therefore, the valuing of the day-by-day cooking should be central in food and nutrition educational actions as an emancipatory and self-care practice [13].

The main source of cooking knowledge and skills is through parents [14; 15; 16; 17]. This information highlights the importance of adult cooking skills as a role model in food preparation habits development in children. In addition, Sidenvall *et al*. (2001) [18] found from a literature review that when changes in household dynamics happen (e.g., when a child moves away from the family or a divorce), the food provider may change their food habits and frequency of meal preparation, which may negatively affect their food choices.

In this scenario, culinary skills among adults, especially those responsible for preparing household meals, have been an important focus of research [14; 17; 19]. Among the publications on this subject are studies that propose to develop instruments that measure culinary skills in adults through the analysis of their psychometric properties.

Before being considered suitable, the instruments must offer accurate, valid, and interpretable data for the population’s assessment. Moreover, the measures are supposed to provide scientifically robust results. These results are established based on measures of reliability and validity of the instruments [20-22]. Reliability is the ability to reproduce a consistent result in time and space or from different observers, demonstrating aspects of stability and internal consistency. It is one of the main quality criteria of an instrument [21]. Validity refers to the fact that a tool measures exactly what it proposes to measure. It is based on extent theory research and experts’ judgement (content validity), the degree in which a group of variables represents the construct to be measured (construct validity) and the degree in which the instrument is related to some external criterion, considered a widely accepted measure (criterion validity) [21-23].

There are public health policies focused on cooking in several parts of the world [3]. Despite the importance of developing instruments that measure culinary skills as a strategy to assist the planning food and nutrition educational actions based on culinary practices, studies have shown lack of methodological uniformity to report validity and reliability of their instruments.

McGowan *et al* (2014) [24] conducted a review of the literature relating to the composition and measurement of an individual’s domestic Cooking Skills (CS) and Food Skills(FS), providing a conceptual and critical analysis of existing measures, and reported on associations of CS and FS with dietary outcomes. However, searches were limited to journal articles in English and limited psychometric data was available in the included studies. Furthermore, the subject of food practices in public health is rapidly evolving, and other culinary skills measurement tools are likely to have been published since they reviewed the literature in 2014.

Additionally, previous reviews have not proposed to appraise the quality of psychometric properties of instruments measuring culinary skills, which justifies the importance of this study, given the fact that the diagnosis of one’s skills entrusted to the application of these instruments may be flawed. This could result in planning inappropriate food and nutrition educational actions for providing emancipatory and self-care practices.

Therefore, this systematic review aimed to identify studies that have developed instruments to measure culinary skills in adult population, and critically appraise the quality of their psychometric properties.

We hope that this study can provide evidence-based guidance on the psychometric properties of instruments measuring culinary skills, to subsidize the selection of valid and reliable instruments by healthcare professionals to assess these subjects in clinical and public health settings and avoid unrealistic expectations about the information that such measures may provide.

## Methods

We registered the protocol of this systematic review on the International Prospective Register of Systematic Reviews (PROSPERO database; http://www.crd.york.ac.uk/PROSPERO/; registration number CRD42019130836). The protocol is available in the S1 Appendix. The PRISMA [25] (Preferred Reporting Items for Systematic Reviews and Meta-Analyses) guidelines for reporting systematic reviews were used to undertake the present review (S1 Table).

### Search strategy

We performed a comprehensive literature search for articles published until January 12, 2021, in the Scopus, LILACS, PubMed, and Web of Science databases. The search strategy included the use of MeSH terms or text words related to the culinary skills, instruments, and validation studies. The PubMed/Medline search strategy was adapted from Terwee, Jansma, Riphagen *et al*. [26]. The S2 Appendix shows the full search strategy for all databases. In addition, we conducted a grey literature search in Google Scholar to identify studies not indexed in the databases listed above. We also evaluated references to the articles found, in order to include any potential studies not yet identified.

### Study selection

This review, included articles meeting the following criteria: 1) address culinary skills in adults; 2) describe the instrument’s validation and reliability process, which can be original or adapted instruments. No filters for year of publication, country or language were employed. Articles that developed original instruments or reporting cross cultural adaptation of instruments addressed to measure culinary skills in children and adolescents or those whose instruments were not available (in the article or upon request to the authors) were excluded. For initial screening of abstracts and titles, we used the *Rayyan Web Platform for Systematic Reviews* [27]. Two authors (A.R.T. and D.B.) independently screened the titles and abstracts of citations to identify potentially relevant studies. We obtained and reviewed the full-text articles for further assessment according to the inclusion and exclusion criteria. When we could not obtain the full text, we contacted the corresponding authors by e-mail or other tools, such as ResearchGate (www.researchgate.net). A third reviewer (T.M.L.) resolved any doubts or disagreements between the reviewers regarding the inclusion or exclusion of articles. The third reviewer compared the results of the independent selection of articles carried out by the two reviewers. If the third reviewer identified any differences, he would ask the two authors to discuss their opinions. If the two reviewers did not reach an agreement, the third reviewer would present his opinion.

### Data extraction and analysis

Two authors (A.R.T. and D.B.) independently performed data extraction using a preformatted spreadsheet in Microsoft Excel. A third reviewer (T.M.L.) resolved any disagreements or doubts resolved any disagreements or doubts occurred in this step, by comparing the data extraction carried out by the two reviewers. If the third reviewer identified any differences, he would ask the two authors to discuss their interpretations. If the two reviewers did not reach an agreement, the third reviewer would present his opinion. We also consulted the third reviewer in case of any doubts regarding the inclusion of potentially relevant articles identified during this step of the systematic review.

The information extracted consisted of descriptive data of the study (country, phenomenon studied, participants, sample size, instrument format, target public, number of items and domains of the instrument, development methodology and statistics performed to report psychometric properties).

### Quality of psychometric properties

We determined the psychometric quality according to the rating system adapted from Hair Jr, Black, Babin *et al*.[28]; Pedrosa, Suárez-Álvarez, and García-Cueto [29]; and Terwee *et al*. (2007) [30]. The criteria addressed the following properties: a) reliability, including internal consistency and stability; b) validity, including content, construct (structural, hypothesis testing and cross-cultural) and criteria. We reported the quality of each measurement property as adequate (+), indeterminate (?), inadequate (-), or no information available (0). When the appraisal of the quality of a specific attribute was not applicable, we reported as ‘NA’. Table 1 shows the quality criteria for psychometric properties. Two independent authors (A.R.T. and D.B.) applied this rating system, and the third reviewer (T.M.L.) resolved any divergences between them (i.e. no consensus on the rating regarding the quality of measure of an instrument), by comparing the results of critical appraisal of the quality of psychometric properties of the instruments, carried out by the two reviewers. If the third reviewer identified any differences, he would ask the two authors to discuss their opinions. If the two reviewers did not reach an agreement, the third reviewer would present his opinion.

**Table 1.**
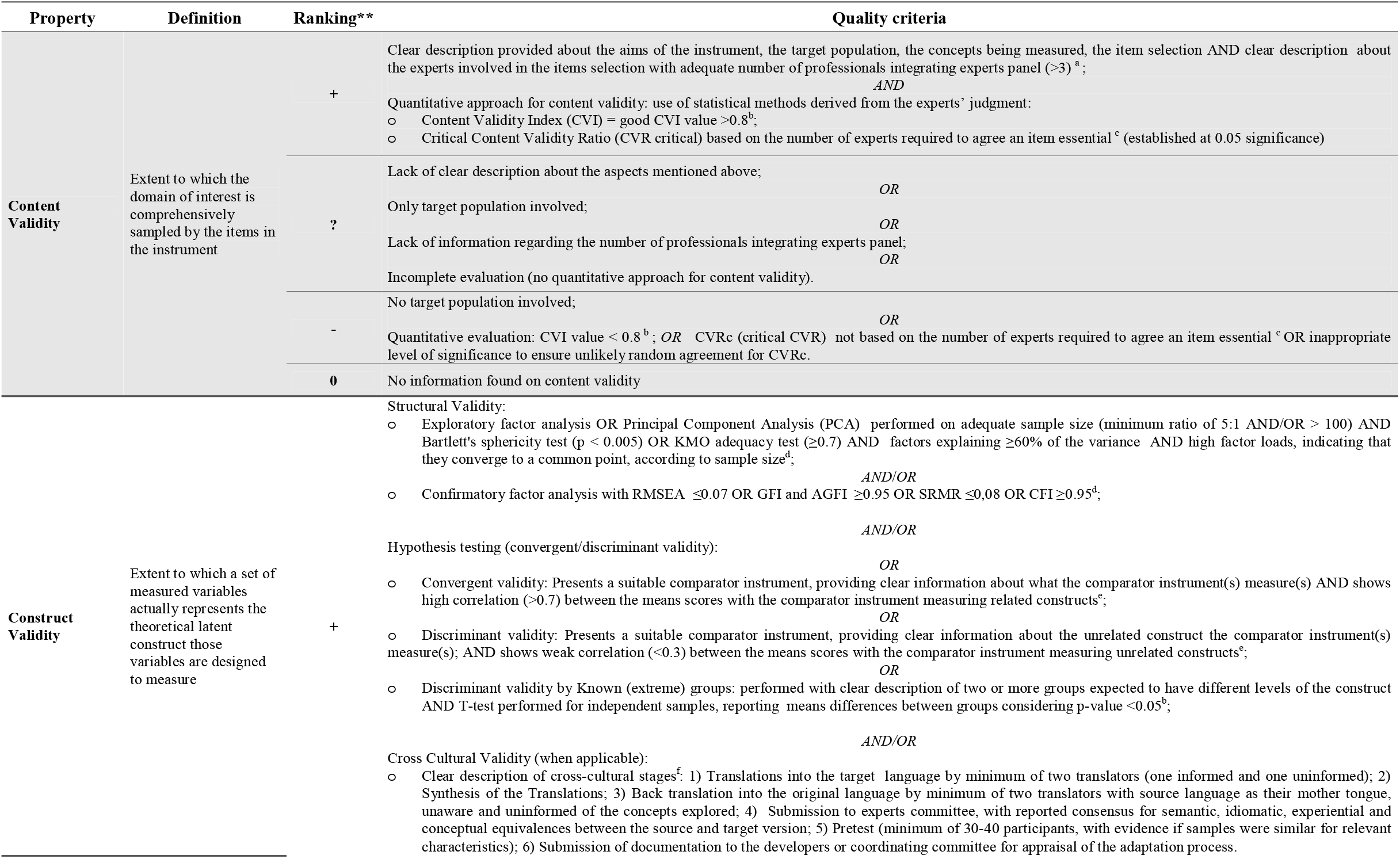
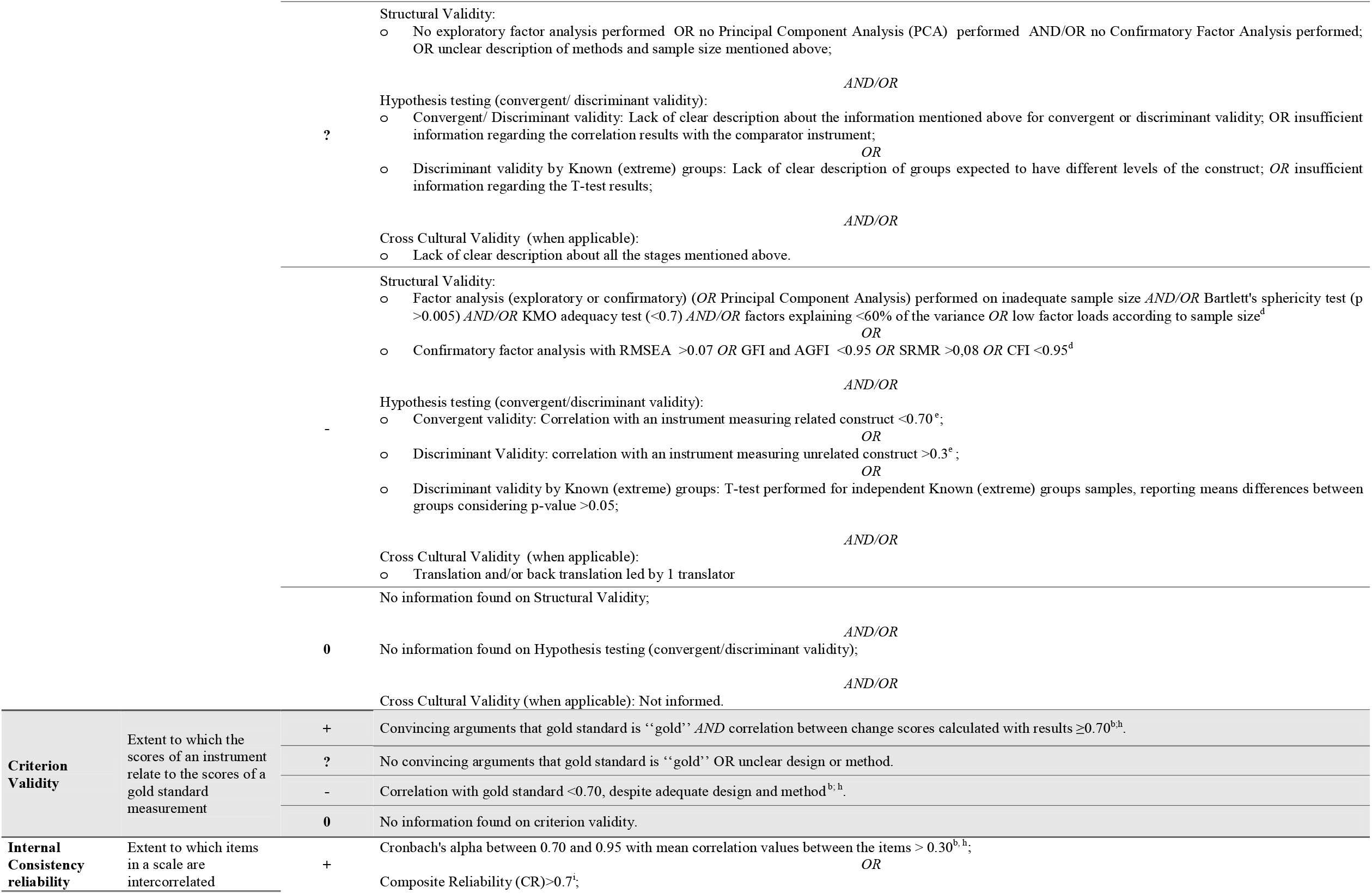
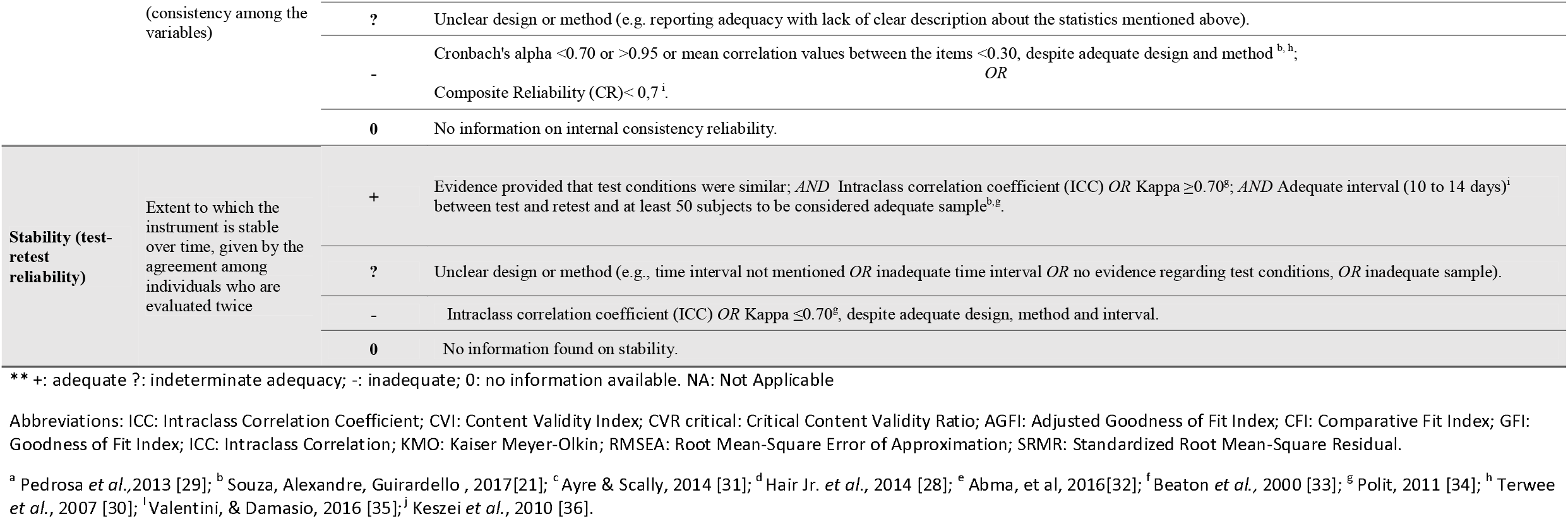
Quality criteria for psychometric properties of measurement (adapted from Hair Jr *et al*. [28], Pedrosa *et al*. [29], and Terwee *et al*. [30]).

## Results

### Search results

The electronic search (including gray literature databases) identified 1148 potentially relevant studies. After reviewing the titles and abstracts, we selected 16 articles for full-text examination. Of these, nine studies (Michaud, 2007 [37]; Warmin, 2009 [38]; Condrasky *et al*., 2011 [39]; Jomori *et al*, 2017 [40]; Barton *et al*, 2011 [41]; Kowalkowska *et al*, 2018 [42]; Lavelle *et al*, 2017 [43]; Kennedy *et al*, 2019 [44]; Martins *et al*, 2019[45]) met the inclusion criteria for review. A list of the excluded studies is available in the S2 Table. The authors presented only one doubt during the selection and data extraction processes, which was resolved by the third reviewer. The doubt corresponded to the inclusion of a potentially relevant article identified during the full text reading of the articles (Hartmann *et al*, 2013[46]). We identified other two relevant studies, by searching the reference lists of the included studies (Vrhovnik, 2012 [47]; Condrasky *et al*, 2013 [48]). Finally, 12 studies were included in this systematic review. Fig. 1 shows a flowchart of the literature search.

**Fig 1.**
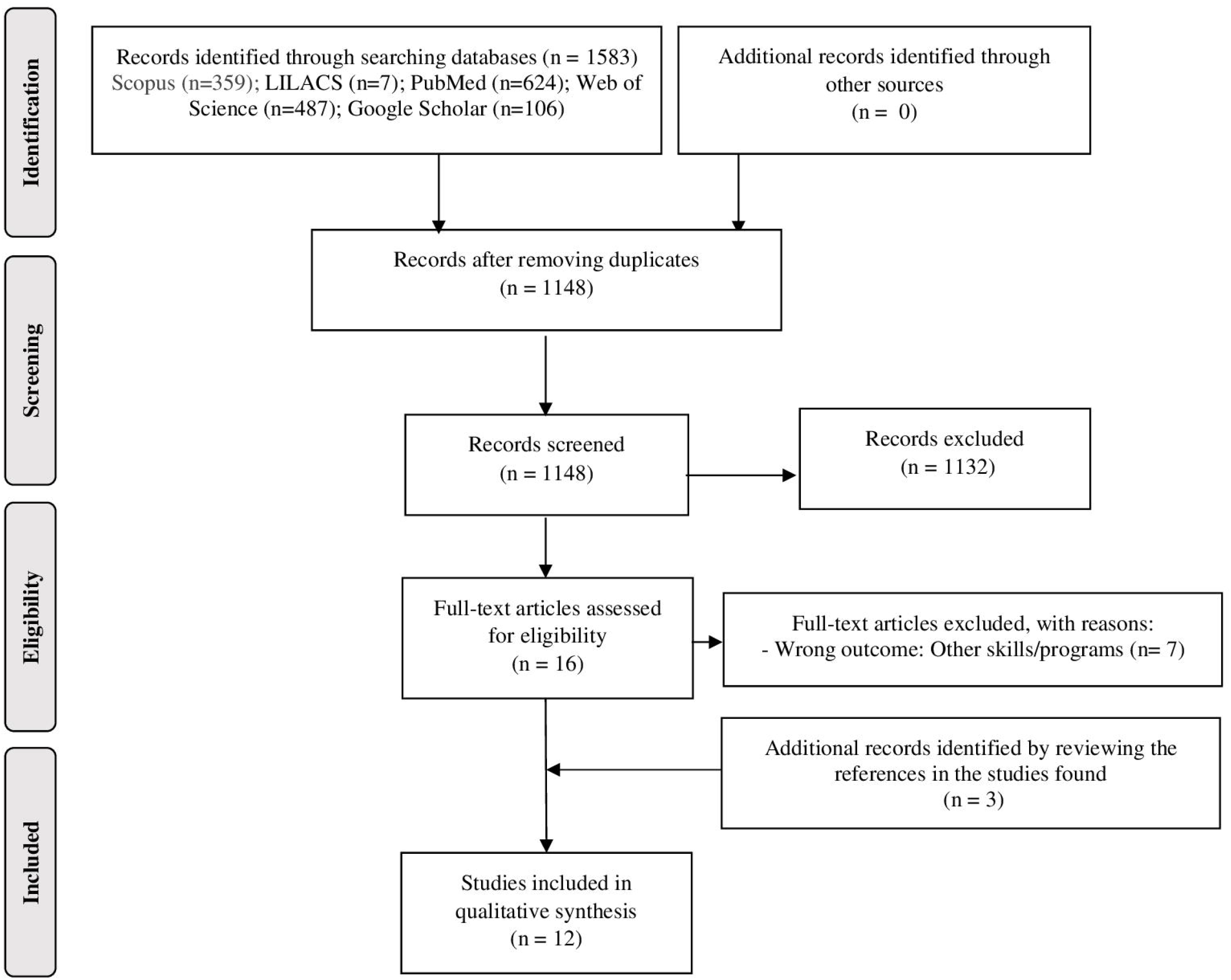
Study selection flowchart of literature search. Abbreviations LILACS: Latin American and Caribbean Health Sciences Literature.

### Characteristics of the studies

Studies were carried out in the United States of America (Michaud, 2007 [37]; Warmin, 2009 [38]; Condrasky *et al*, 2011 [39]; Condrasky *et al*, 2013 [48]), Brazil (Jomori *et al*, 2017 [40]; Martins *et al*, 2019 [45]), Canada (Vrhovnik, 2012 [47]; Kennedy *et al*, 2019 [44]), Switzerland (Hartmann *et al*, 2013 [46]), Portugal (Kowalkowska *et al*, 2018 [42]), Scotland (Barton *et al*, 2011 [41]) and Northern Ireland and Republic of Ireland (Lavelle *et al*, 2017 [43]). All of them were published in English, between 2007 and 2019. One study did not seek ethical approval (Barton *et al*, 2011 [41]).

Included papers had distinct purposes: those reporting the development of an original instrument, or cross-cultural adaptation of a tool to explicitly measure cooking/food skills or a part thereof (n = 7) and original tools developed to evaluate a cooking and food skills intervention (n = 5). Most tools assessed cooking skills in adults from a particular country (Hartmann *et al*, 2013 [46]; Lavelle *et al*, 2017 [43]; Vrhovnik, 2012 [47]; Kennedy *et al* l, 2019 [44]), parents of schoolchildren responsible for food preparation at home (Martins *et al*, 2019 [45]), university students (Warmin, 2009 [38]; Jomori *et al*, 2017 [40]; Kowalkowska *et al*, 2018 [42]) and adults of low-income communities, participants in culinary skills and nutrition education programs (Michaud, 2007 [37]; Condrasky *et al*, 2011 [39]; Condrasky *et al*, 2013 [48]; Barton *et al*, 2011 [41]). Study samples were mostly composed of women (Barton *et al*, 2011 [41], Condrasky *et al*, 2011 [39]; Martins *et al*, 2019 [45]; Kennedy *et al*, 2019 [44]; Kowalkowska *et al*, 2018 [42]; Michaud, 2007 [37]; Warmin, 2009 [38]; Vrhovnik, 2012 [47]). The participants’ age ranged from 18 to 69 years. Sample sizes ranged from 29 to 4.4306 individuals.

Table 2 shows the characteristics of the included studies.

**Table 2.** Characteristics of the included studies.

| Author/year/<br>Country | Objective | Instrument<br>(mnemonic) | Latent phenomenon<br>evaluated | Study sample | Age<br>years/ range<br>/mean(SD) | N |
| --- | --- | --- | --- | --- | --- | --- |
| **Michaud /**<br>**2007 /USA** | To provide evidence for and demonstrate the processes used to develop and test tools to measure the effectiveness of a culinary and nutrition education program | Cooking and healthy eating evaluation questionnaire | Cooking skills and healthy eating, based on the main objectives of the Cooking with a Chef (CWC) program | Experts panel: professionals (nutrition, public health, gastronomy, sociology, statistic);<br>Pilot data and larger study: Parents and caregivers recruited from preschool, public school, church, and organized playgroup settings in South Carolina. | Pilot and larger study: 18 to 50 years old. | Experts panel (n=12);<br>Pilot data (n = 39), with test-retest subgroup (n = 19); Larger study data (n = 162). |
| **Warmin /**<br>**2009 / USA,**<br>**article**<br>**published in**<br>**2012** | To test the effects of an established culinary nutrition program with college students and to test the effectiveness of placing of the nutrition component onto an online presentation. | Cooking and healthy eating evaluation questionnaire (online version; new items) | Cooking skills and healthy eating, based on the main objectives of the Cooking with a Chef (CWC) program | College students recruited from a Nutrition for non-majors class offered through Clemson University's Food Science and Human Nutrition Department, who received no 'Cooking with a Chef' intervention. | 18 to 22 years old. | Test-retest (n=29) |
| **Condrasky et**<br>**al. / 2011 /**<br>**USA** | To develop scales to assess the impact of the Cooking with a Chef program on several psychosocial constructs | Cooking and healthy eating evaluation questionnaire (short version) | Cooking skills and healthy eating, based on the main objectives of the Cooking with a Chef (CWC) program | Experts panel: Academic professionals;<br>Pilot and larger study: Parents, caregivers and cooks, largely female recruited from child care settings in South Carolina and church and school kitchens. | Pilot and larger study: 35 years old or older. | Experts panel (n=4);<br>Pilot data (n = 39), with test-retest subgroup (n = 19); Larger study data from self-selected parents and caregivers (n = 162) and cooks (n=83); |
| **Condrasky et**<br>**al. / 2013 /**<br>**USA** | To develop and evaluate a participatory training for cooks in African American churches. | Cooking and healthy eating evaluation questionnaire (new items) | Cooking skills and healthy eating, based on the main objectives of The Faith, Activity, and Nutrition (FAN) program , adapted from the CWC program | Experts panel: Nutrition professionals;<br>Pilot data: cooks and planning committee members. | Not specified. | Not specified. |
| **Jomori et al.**<br>**/2017 / Brazil** | To describe the results of the construct validity by the known-groups' method of a Brazilian cooking skills and healthy eating questionnaire | Brazilian version of the Cooking and healthy eating evaluation questionnaire | Cooking skills and healthy eating | Students who had started their undergraduate program at Federal University of Santa Catarina (UFSC), Brazil in 2015 were selected based on convenience. They voluntarily accessed the online questionnaire from August to November 2015. | 20.7 (±5.59) years old. | University students (n=767). |
| **Hartmann et** | To design a | Cooking | Cooking skills | Adult participants from the Swiss Food Panel, a | 55.5 years (SD = 14.6, | test-retest: n= 4436. |
| **al / 2013 / Switzerland** | cooking skill scale that is reliable and applicable to most people | Skills Scale (CSS) |  | population-based longitudinal study of the eating behavior of the Swiss population. | range 21–99). |  |
| **Kowalkowska et al. /2018 /Portugal** | To assess the reliability of a Portuguese version of the Cooking Skills Scale (CSS) and to evaluate the association between cooking skills and socio-demographic, psychological and other cooking related variables | Portuguese version of the Cooking Skills Scale (CSS) | Cooking skills | Portuguese university students, attending bachelor's or integrated master's degree studies, with access to cooking facilities. | 22.8 years (SD= 4.9; range: 17 - 58). | Larger study data (n=730);<br>Repeatability - test-retest (n= 106). |
| **Barton et al. / 2011 / Scotland** | To undertake an assessment of validity and reliability of a short questionnaire designed to measure the impact of cooking skills interventions on cooking confidence, the use of basic food skills, and food selections amongst low income communities | Short Questionnaire - CookWell programme | Cooking confidence and food skills, based on the key domains shown to be influenced by the CookWell programme | Experts panel: dietitians and community workers;<br>Face validity: adults residing in Tayside, Scotland, who were typical of those who may attend cooking skills classes;<br>Reliability: group of adults attending community-based classes (other than cooking) in Tayside, Scotland;<br>Feasibility: from participants from the 'Get Cooking' project, living in areas in the lower deciles (most deprived) of the Scottish Index of Multiple Deprivation. | Face validity: range of 21-69 years;<br>Reliability: 46 (15.1);<br>Feasibility: 35.0 (20.8). | Experts panel (n= 28);<br>Face validity (n=20);<br>Reliability (n=57);<br>Feasibility (n =24). |
| **Vrhovnik / 2012/ Canada** | To create a valid and reliable tool to assess the level of food skills in the community | Food skills survey tool | Food skills | Face validity: public health dieticians and nurses;<br>Content validity: Field experts in food skills and survey development from Queen's University, the research department at KFL&A (Kingston, Frontenac, Lennox & Addington Health Unit);<br>Pilot, factor validity and reliability: adults from Kingston, Frontenac, Lennox and Addington counties, able to understand English, recruited through the directories of residential phone numbers provided by CCI Research. | Pilot, factor validity and reliability: age>18 years old. | Face validity (n=13);<br>Content validity (n=10);<br>Pilot, factor validity and reliability (n=- 273). |
| **Lavelle et al. / 2017 / Ireland** | To develop and validate a measure for cooking skills and one for food skills, that are clearly described, relatable, user-friendly, | Cooking skills confidence measure, and Food skills | Cooking skills and food skills | Experts' opinion: professionals working in the area of health promotion including cooking and food skills interventions and education;<br>Study 1: adults responsible for preparing a main | Study 1: 20–60 years old;<br>Study 2: 18 -27 years old; | Study 1 - content and convergent validity (n=1049);<br>Study 2 - test-retest |
|  | suitable for different types of studies, and applicable across all sociodemographic levels. |  |  | meal at least once per week;<br>Study 2: students from Ulster University enrolled on a course that consisted of nutrition, hospitality, food marketing or food product and innovation orientated modules;<br>Study 3: students from the Ulster University, Northern Ireland and St. Angela's College Sligo, Ireland, either studying a Business-related degree or were studying Home Economics, classified as 'Food preparation novices' and 'Experienced food preparers';<br>Study 4: combination of the samples in study 2 and study 3 (representing the P/P method) and participants randomly selected from the sample in Study 1 ((representing the CAPI method). | Study 3: 'Experienced food preparers': 18–26 years old; 'Food preparation novices': 19–24 years old. | and internal consistency reliability of the measures in the P/P format (n = 23);<br>Study 3 - discriminant validity and further assess the internal consistency reliability of the P/P measures (n = 57);<br>Study 4 - differences between the CAPI and the P/P method in relation to the confidence scores of the measure (n= Studies 2 and 3 + 38 from study 1). |
| **Kennedy et al / 2019/ Canada** | To develop, validate, and assess reliability of a food skills questionnaire | Food skills questionnaire | Basic to intermediate food skills | Content validity: Dietitians, home economists, academics, and chefs;<br>Face validity: convenience sample of students at Western University;<br>Test-retest and inter-item reliability: undergraduate students randomly selected. | Test-retest and item reliability: (mean age, 22 ± 6 years. | Content validity (n=17);<br>Face validity (n=20)<br>Test-retest and inter-item reliability (n=165; lowest number of participants answering the same questions at times 1 and 2 was 126). |
| **Martins et al / 2019/ Brazil** | To describe the development and the reliability assessment of an index that evaluates the confidence in performing cooking skills considered relevant in Brazil. | Cooking Skills Index (CSI) | Cooking skills self-efficacy | Face validity and experts panel: nutritionists, physicians and biologists belonging to the research group that supported the preparation of the Dietary Guidelines for the Brazilian Population;<br>Pilot, test- retest and internal consistency: parents of schoolchildren responsible for food preparation at home. | Not specified. | Face validity and experts panel (n= 6);<br>Pilot (n=10) and test-retest and internal consistency (n=51). |

### Characteristics of the instruments

All studies provided description of the construct, with conceptual framework or clear rationale to define their instruments’ construct.

Six studies reported the development (Michaud, 2007 [37]; Warmin, 2009 [38]; Condrasky *et al*, 2011 [39]; Condrasky *et al*, 2013 [48]; Barton *et al*, 2011 [41]) or cross-cultural adaptation of tools (Jomori *et al*., 2017 [40]) aiming to evaluate cooking skills and healthy eating, based on the main objectives of cooking and nutrition education interventions programs.

Michaud (2007) [37], developed an original questionnaire, consisting of 51 items measuring culinary skills and healthy eating, aiming to evaluate the Cooking with Chef (CWC) intervention Program. In 2009, Warmin [39] tested the online format of application of this questionnaire, based on a sample of university students. In addition, Condrasky *et al*. (2011) [39] reported the alteration of three scales in this questionnaire. Condrasky *et al*. (2013) [48] then adapted a few items of the questionnaire, employed in a sample of church cooks in South Carolina (USA). The final questionnaire consisted of 64 items, with one knowledge evaluation section, a short index and six scales related to Self-Efficacy for produce consumption, cooking, using basic techniques, using fruits, vegetables, and seasonings during cooking practices. Finally, Jomori *et al*. (2017) [40] described the results of a cross-cultural adaptation of the later version of the culinary skills and healthy eating questionnaire for Brazilian students and reported its construct validity.

Barton *et al*. (2011) [41] also described the results of the development and validation of a short cooking skills questionnaire, aiming to evaluate the effects of the CookWell intervention program. The questionnaire consisted of 19 items with domains related to frequency of preparing food, confidence in following a simple recipe, cooking with basic ingredients, and preparing new foods and recipes. Some items were similar to those reported in the cooking skills and healthy eating questionnaire, based on the main objectives of the Cooking with a Chef (CWC) program, described in the aforementioned studies (Michaud, 2007 [37]; Warmin, 2009 [38]; Condrasky *et al*., 2011 [39]; Condrasky *et al*., 2013 [48]; Jomori *et al*., 2017 [40]).

The remaining studies (Hartmann *et al*., 2013 [46]; Kowalkowska *et al*., 2018 [42]; Lavelle *et al*., 2017 [43]; Vrhovnik, 2012 [47]; Kennedy *et al*., 2019 [44]; Martins, *et al*.al, 2019 [45]) described the results of the development and validation or cross-cultural adaptation of tools aiming to evaluate adults’ cooking and/or food skills or a part thereof, with some similarities. The instruments’ domains ranged from 1 to 3 and the number of items ranged from 7 to 39, mainly related to ‘food preparation techniques’, ‘meal planning’ and ‘food selection and purchase’. We present the main characteristics of these instruments, their domains and items in common as well as the divergences below.

The Cooking Skills Scale (originally developed by Hartmann *et al*., 2013 [46]; and adapted for Portuguese university students, by Kowalkowska *et al*., 2018 [42]) focused on measuring cooking skills, based on the ability to prepare certain dishes and products, but without distinguishing whether they were prepared with basic ingredients, pre-prepared products, convenience foods or a combination of them.

Lavelle *et al*. (2017) [43] developed measurements to assess cooking skills and food skills confidence. The cooking skills confidence measure consisted of 2 domains: ‘Food preparation Techniques’ and ‘Cooking method’, including items related to skills for cooking pre-prepared products and convenience foods (e.g.: *rate how good they are at: Microwave food, including heating ready-meals*).

Unlike the items shown in Hartmann *et al*.’s cooking scale (2013) [46] and Lavelle *et al*.’s cooking confidence measure (2017) [43], the Cooking Skills Index, developed by Martins, *et al*., (2019) [45] focused on cooking self-efficacy related to the preparation of meals from the combination of natural or minimally processed foods and seasoned using natural seasonings and culinary ingredients.

Lavelle *et al*.’s food skills confidence measure (2017) [43] consisted of five domains related to meal planning, shopping, budgeting, resourcefulness and label reading. Kennedy *et al*.*’*s food skills questionnaire (2019) [44] focused on similar domains, such as ‘Food Selection and Planning’, ‘Food Safety and Storage’; however, it comprised one domain related to ‘Food Preparation’. Like Martins *et al*.’s [45] instrument for accessing cooking skills (2019), this domain included items to assess confidence in performing cooking techniques, (e.g.: *rate your confidence in boiling, steaming or stewing*) and using basic ingredients and seasoning (e.g.: *rate your confidence in: preparing food from basic ingredients; choosing a spice or herb*). Vrhovnik’s Food skills survey tool (2012) [47] also consisted of domains regarding ‘Mechanical Techniques’ (using texture, taste and smell to guide cooking methods), ‘Food Preparation’ (chopping, mixing, blending, cooking and following recipe) and ‘Conceptualizing Foods’ (creating meal ideas with leftover food and adjusting recipes to fit the needs of an individual).

The studies reported analysis of the psychometric properties of their instruments: Only two studies presented statistical methods derived from the experts’ judgment for content validity (Kennedy *et al*., 2019 [44], Vrhovnik, 2012[47]). Six out of ten studies that proposed to develop and validate a new instrument (or those reporting small changes in the original tool) did not report construct validity (Warmin, 2009 [38]; Condrasky *et al*., 2013 [48], Hartmann *et al*., 2013 [46]; Barton *et al*., 2011[41]; Kennedy *et al*., 2019 [44]; Martins *et al*., 2019 [45]). Two studies reported cross-cultural adaptation of instruments (Kowalkowska *et al*., 2018 [42], Jomori *et al*., 2017 [40]). Only one study (Michaud, 2007 [37]) reported criterion validity. Nine studies tested the reliability of their instrument according to internal consistency (Michaud, 2007 [37]; Condrasky *et al*., 2011 [39]; Jomori *et al*., 2017 [40]; Kowalkowska *et al*., 2018 [42]; Barton *et al*., 2011 [41]; Vrhovnik, 2012 [47]; Lavelle *et al*., 2017 [43], Kennedy *et al*., 2019 [44]; Martins *et al*., 2019 [45]) and/or stability (Michaud, 2007 [37]; Warmin, 2009 [38]; Condrasky *et al*., 2011 [39]; Barton *et al*., 2011 [41]; Kowalkowska *et al*., 2018 [42]; Lavelle *et al*., 2017 [43], Kennedy *et al*., 2019 [44]; Martins *et al*., 2019 [45]; Hartmann *et al*., 2013 [46]). Table 3 describes the characteristics of the instruments.

**Table 3.**
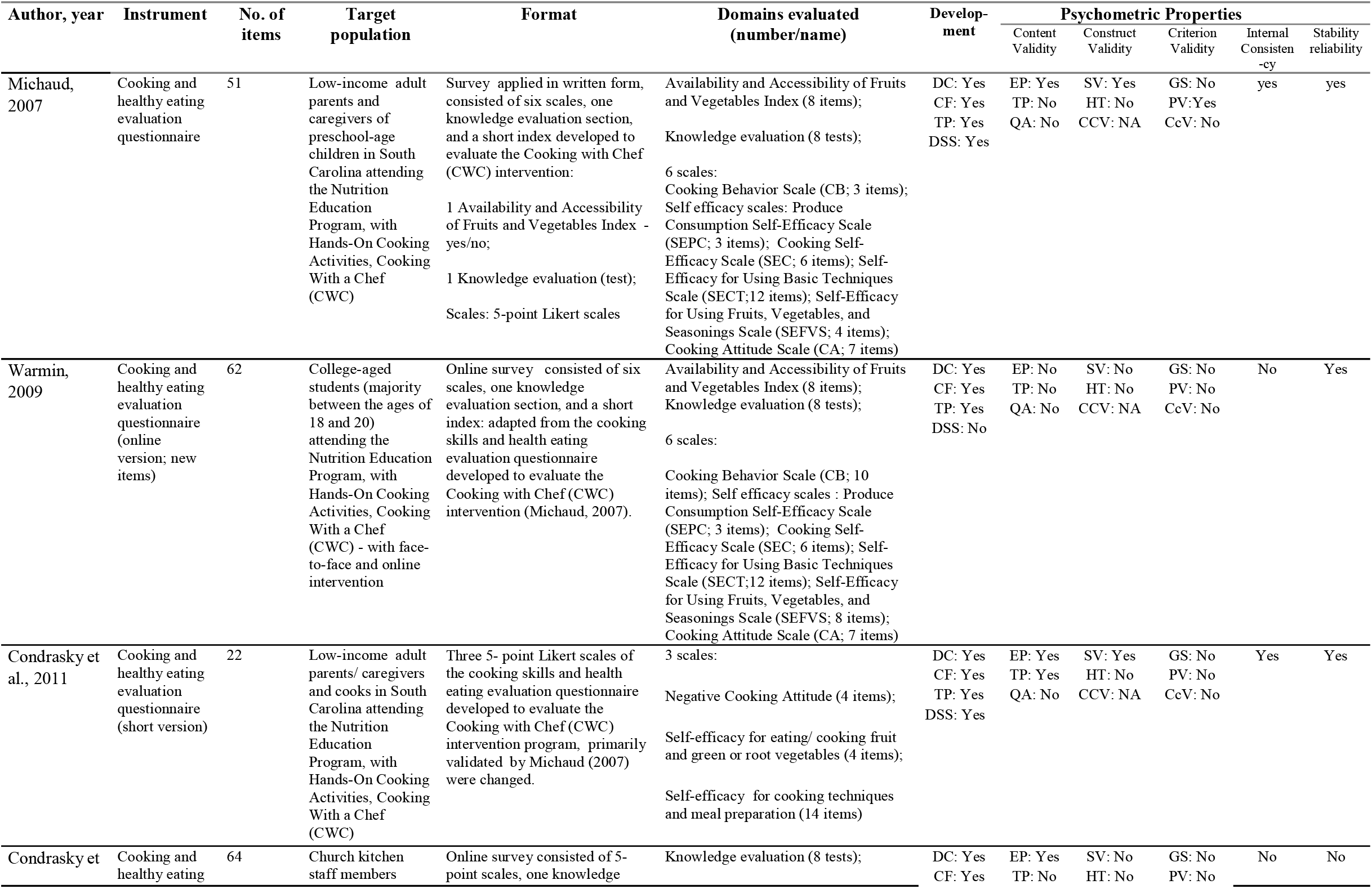
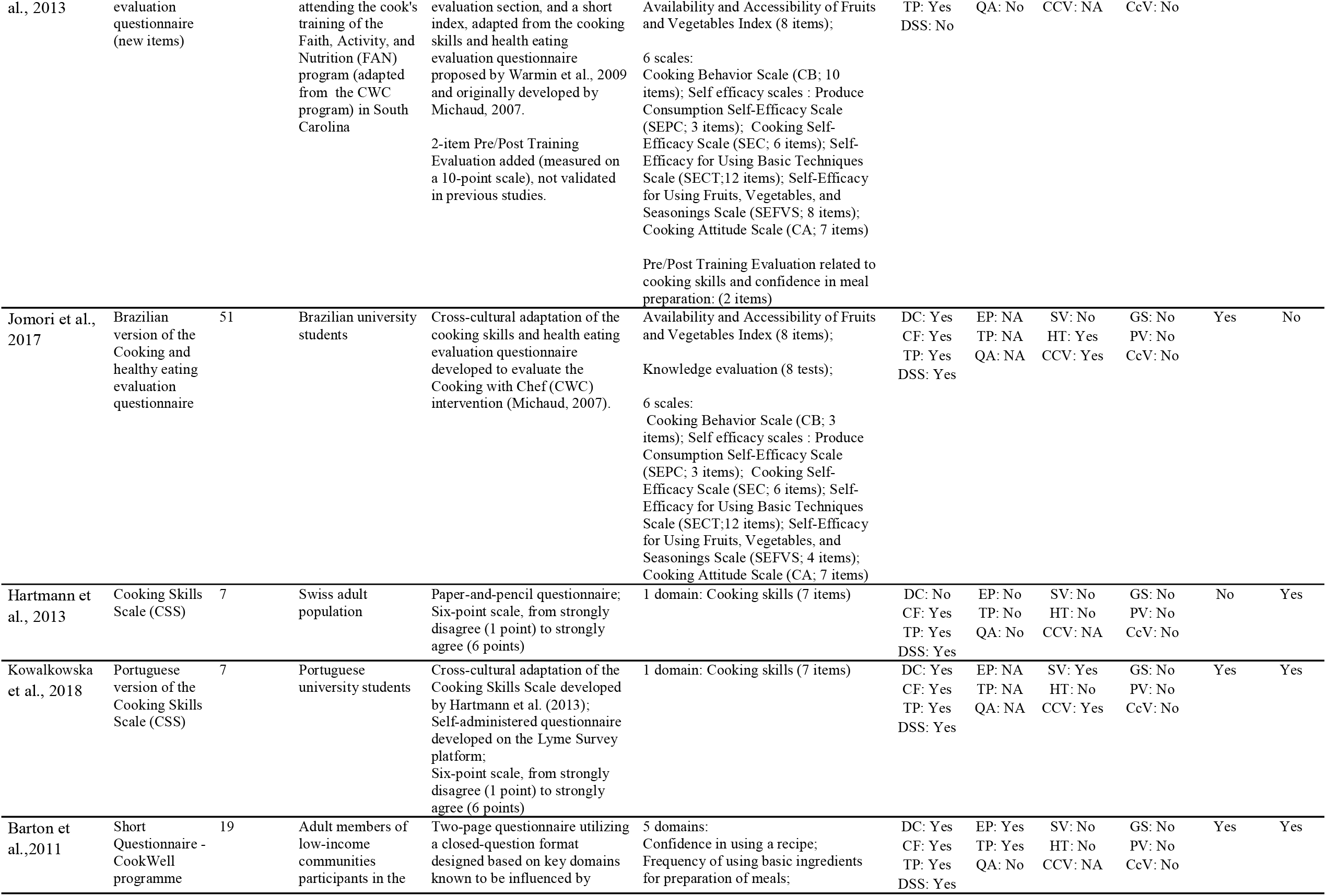
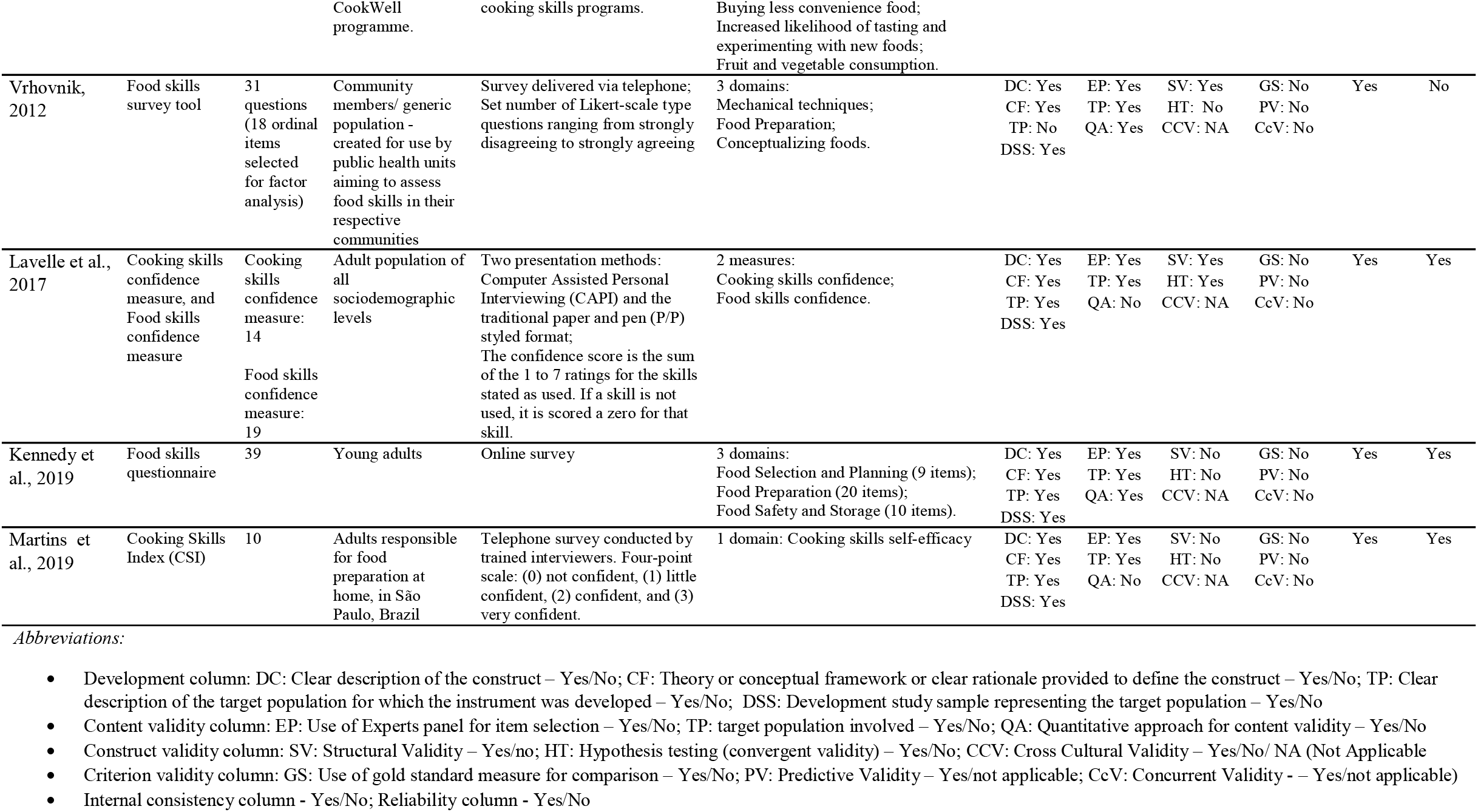
Characteristic of the instruments.

### Quality of the psychometric properties

We describe the quality of the psychometric properties of the instruments in Table 4.

**Table 4.**
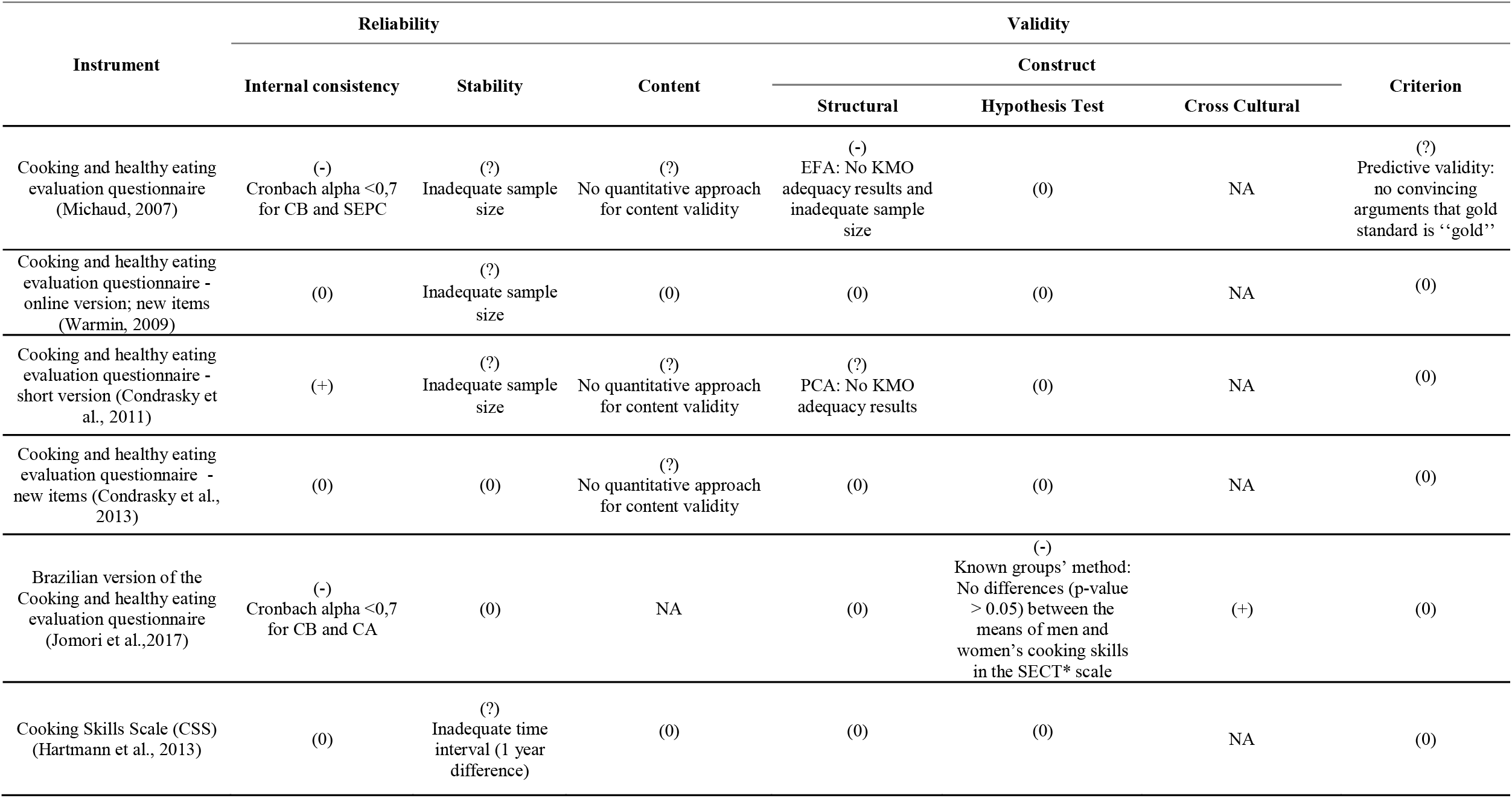
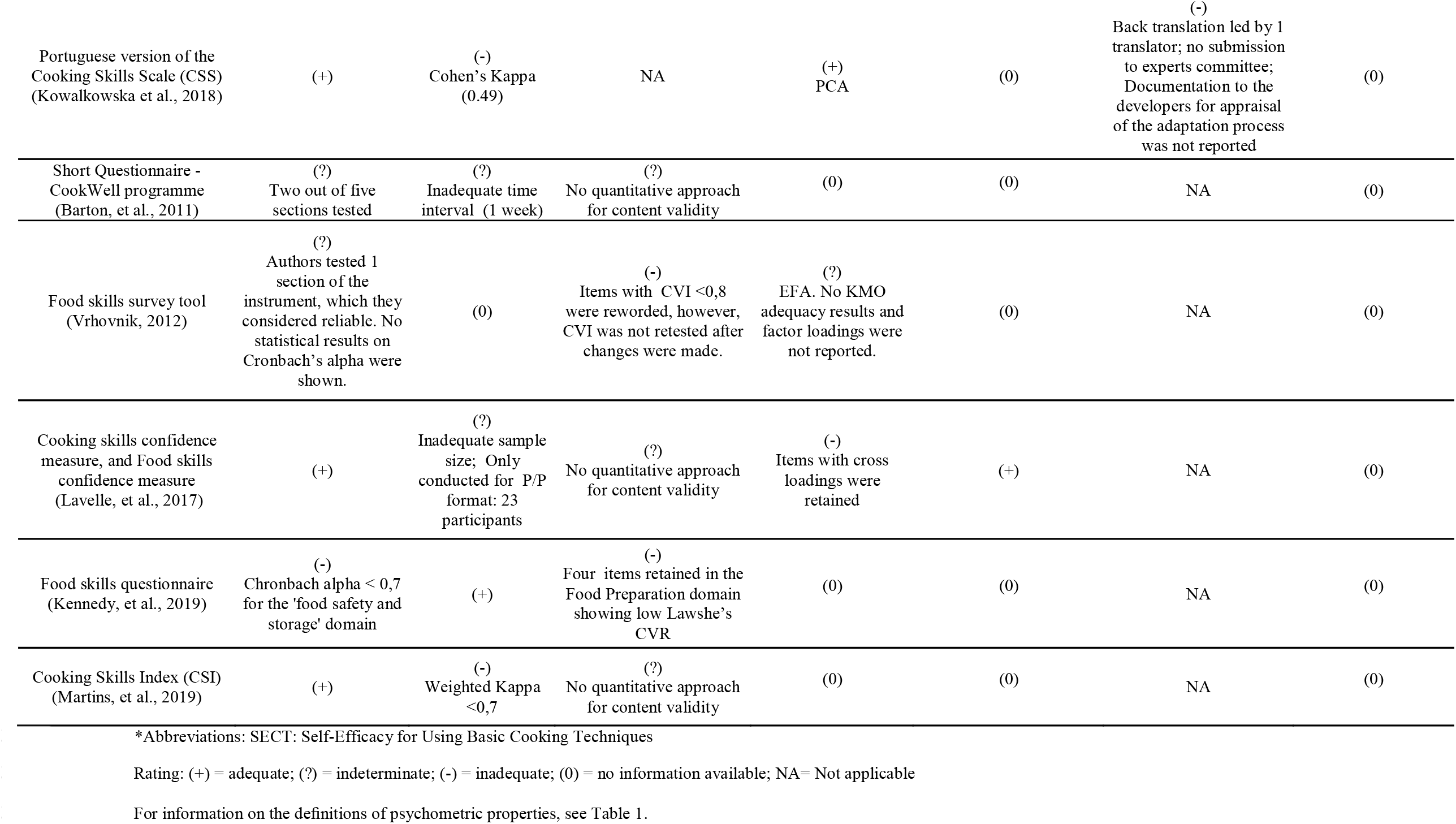
Evaluation of quality of psychometric properties of the instruments.

We considered adequate quality of internal consistency reliability in four studies (Condrasky *et al*., 2011 [39]; Kowalkowska *et al*., 2018 [42]; Lavelle *et al*., 2017 [43]; Martins *et al*., 2019 [45]) Three studies (Michaud, 2007 [37]; Jomori *et al*., 2017 [40]; Kennedy *et al*., 2019 [44]) showed inadequate quality of this measure. Two studies had the internal consistency reliability considered indeterminate: Barton *et al*., 2011 [41] did not test three out of five sections of their instrument for internal consistency reliability; Vrhovnik, 2012 [47] did not report any statistical results, despite the author’s affirmation on satisfactory results for internal consistency. Three studies (Warmin, 2009 [38]; Condrasky *et al*., 2013 [48]; and Hartmann *et al*., 2013 [46]) did not report internal consistency reliability.

Nine studies reported test-retest reliability (Michaud, 2007 [37]; Warmin, 2009 [38] Condrasky *et al*., 2011 [39]; Hartmann *et al*., 2013 [46]; Kowalkowska *et al*., 2018 [42]; Barton *et al*., 2011 [41]; Lavelle *et al*., 2017 [43], Kennedy *et al*., 2019 [44]; and Martins *et al*., 2019 [45]). However, we considered the quality of this measure inadequate in two of these studies (Kowalkowska *et al*., 2018 [42]; Martins *et al*., 2019 [45]), since they presented results inferior to the minimum criterion for *Kappa*, despite adequate design and method. Five studies showed inadequate time interval (Barton *et al*., 2011 [41]; Hartmann *et al*., 2013 [46]) or inadequate sample size for test-retest reliability analysis (Michaud, 2007[37]; Warmin, 2009 [38]; Condrasky *et al*., 2011 [39]; Lavelle et. al, 2017 [43]); therefore, we deemed the quality of stability inadequate in these studies.

No studies reporting the development or small changes of an original instrument provided adequate measures to show content validity. The authors did not calculate any index of agreement for content validity (Martins *et al*., 2019 [45]; Lavelle *et al*., 2017 [43]; Barton *et al*., 2011 [41], Condrasky *et al*., 2011 [39]; Condrasky *et al*., 2013 [48]; Michaud, 2007 [37]), or statistical results of expert’s agreement did not reach minimum criteria to be considered valid (Vrhovnik, 2012 [47]; Kennedy *et al*., 2019 [44]). Moreover, one study did not perform any analysis of content validity (Hartmann *et al*., 2013 [46]).

Regarding construct validity, six studies reported at least one kind of analysis (structural validity, hypothesis testing or cross-cultural validity, when applicable).

Five studies performed structural validity analysis (Michaud, 2007 [37]; Condrasky *et al*., 2011 [39]; Vrhovnik, 2012 [47]; Kowalkowska *et al*., 2018 [42]; Lavelle *et al*., 2017 [43]). We classified two of them as inadequate according to the quality of this attribute, due to insufficient sample size for Exploratory Factor Analysis (EFA) (Michaud, 2007 [37]) and retention of items showing cross loadings (Lavelle *et al*., 2017 [43]). In addition, one study performed exploratory factor analysis, however did not provide results for factor loadings (Vrhovnik, 2012 [47]), hence, we considered indeterminate quality of structural validity. Only two studies reported hypothesis testing for construct validity. One of them properly performed convergent validity with satisfactory results (Lavelle *et al*., 2017 [43]). Jomori *et al*., 2017 [40] performed discriminant validity between known groups, however we rated the quality of this attribute inadequate, since the authors reported no significant differences between groups in one scale (*Self-Efficacy for Using Basic Cooking Techniques (SECT)*). Two studies performed cross-cultural adaptations of original instruments (Jomori *et al*., 2017 [40] and Kowalkowska *et al*., 2018 [42]). One of them received inadequate rating due to insufficient number of translators leading back translation, and stages for cross-cultural adaptation were incomplete (Kowalkowska *et al*., 2018 [42]). Six studies did not report any kind of analysis to evidence construct validity (Warmin, 2009 [39]; Condrasky *et al*., 2013 [48]; Hartmann *et al*., 2013 [46]; Barton *et al*., 2011 [41]; Kennedy *et al*., 2019 [44]; Martins *et al*., 2019 [45]).

Most studies did not provide information on criterion validity. Only one study (Michaud, 2007 [37]) performed analysis for criterion validity, however, the authors did not describe it clearly (convincing arguments for gold standard).

## Discussion

### Summary of evidence

To our knowledge, this is the first systematic review to identify and appraise quality of psychometric properties of instruments for assessing culinary skills in adults. This article has provided a comprehensive critical analysis of the studies’ characteristics and their psychometric properties. We found twelve studies developing original instruments to measure culinary skills in adults, or performing cross-cultural adaptations.

This systematic review has highlighted gaps in these instruments, suggesting the need to develop new studies with robust and standardized psychometric methodology that shows validity and reliability of culinary skills measurements. Although we considered adequate quality of internal consistency reliability in four studies, only one study received adequate rating for stability (test-retest reliability). No studies developing original instruments presented satisfactory measurement for content validity since the authors did not calculate any index of agreement. Only four studies showed satisfactory results for at least one type of construct validity (structural, hypothesis testing or cross-cultural adaptation, when applicable) and only one study reported criterion validity, however, we considered inadequate quality of this measurement property. These results indicate that although there are isolated measures appraised in this review that show good promise in terms of quality of psychometric properties, no studies presented satisfactory results for each aspects of reliability and validity.

### General view of the studies

Most studies are originally from countries whose native language is English. One Brazilian study (Martins *et al*., 2019 [45]) originally developed an instrument in Portuguese for application with parents of schoolchildren, responsible for food preparation at home. However, the authors translated the instrument from Brazilian Portuguese into English, without making it available in its original language. Despite the authors’ intention to provide access to their study using universal language, translating the instrument into English is not enough to guarantee its international applicability, considering cultural aspects. Developing a new instrument in one’s own language or adapting existing instruments to each setting is necessary to guarantee the instruments’ linguistic and cultural appropriateness [33].

Regarding submission of psychometric studies for ethical approval, one study (Barton *et al*., 2011 [41]) justified the absence of ethical approval because it comprised developmental work for service evaluation. Despite the fact that validation studies aim at the development of tools for measuring latent phenomena, methods applied to report the reliability and validity of such instruments involve the participation of human beings; therefore, the submission of such studies to ethical approval is not only essential, but also indispensable [49; 50].

### General view of the instruments

Most studies reported the development of scales, indexes, and questionnaires. One study classified their instrument as an index (Martins *et al*., 2019 [45]); however, the instrument used Likert scale to register participants’ statements related to the assessed latent phenomenon (cooking self-efficacy). It is important to highlight differences between an index and a scale. An index compiles one score from an aggregation of two or more indicators that attempt to signal, by means of a value, both a content relation with the represented phenomenon and the evolution of a quantity in relation to a reference. The indicator communicates or reveals progress toward a certain goal, and it is applied as a resource to make a tendency or phenomenon not immediately detectable by isolated data more noticeable. It represents an essential tool for the decision-making process and social control, and it is not an explanatory or descriptive element, but provides punctual information on time and space, whose integration and evolution can activate or accompany reality [51].

A scale, on the other hand, measures levels of intensity at the variable level, like to what extent a person agrees or disagrees with a particular statement. A scale is a type of measure composed of several items that have a logical or empirical structure among them. The most commonly used scale is the Likert scale. The sum of scores for each of the statements creates an overall score of the intensity related to the assessed latent phenomenon [20].

The majority of the included studies presented instruments with items assessing cooking self-efficacy (regarding food preparation techniques), meal planning and food selection and purchase. The main difference between the instruments referred to the conceptualization of culinary skills: some authors comprehend that such skills comprise the ability to prepare certain dishes, including those based on pre-prepared products and convenience foods (Hartmann *et al*., 2013 [46]; Kowalkowska *et al*., 2018 [42]; Lavelle *et al*., 2019 [43]). However, relying on pre-prepared or convenience products to prepare a meal may require less cooking abilities [10]. Thus, using the microwave oven for the mere heating of frozen meals, for example, could overestimate an individual’s skill level. Moreover, pre-prepared products and convenience foods are often classified as ultra-processed foods, whose negative nutritional attributes are associated with harmful effects to health [52]. Hence, subsidizing the choice of instruments that enable the assessment of culinary skills and healthy culinary practices, based on the aforementioned domains, is essential for Public Health scenario.

Some authors identify cooking skills as a distinct construct from food skills. Lavelle *et al*. (2017) [43] define cooking skills as a set of physical or mechanical skills used in the production of a meal while food skills are described as a wider set of skills involved in the entirety of the meal preparation process that includes meal planning, shopping, budgeting, resourcefulness, and label reading. Short [6] however, specifies that reducing cooking skills to the ability to do tasks such as baking, broiling, poaching, and stir-frying is an oversimplification of activities involved in planning, organizing, and preparing a meal. She also states that our confidence in cooking and using basic skills influences what and how we cook, which may influence our diet quality. Kennedy *et al*. (2019) [44] seem to consider mechanical skills for meal production as part of the overall construct of food skills. The authors also state that low food skills or cooking self-efficacy are barriers to healthy eating. Vrhovnik (2012) [47] conceptualizes food skills as necessary abilities for knowledge, planning, conceptualization, preparation and perception of food. Although these authors quoted such domains to define the construct of food skills, they seem to be aligned with the concept of cooking skills adopted in this review [7], reinforced by Short [6].

### Psychometric quality

Although all instruments reported some psychometric information, the evaluation of the psychometric quality using the criteria adopted in this systematic review exhibited some missing data.

Regarding the reliability of the instruments, most studies reported internal consistency reliability (Michaud, 2007 [37]; Condrasky *et al*.., 2011 [39]; Jomori *et al*., 2017 [40], Kowalkowska *et al*., 2018 [42], Barton *et al*., 2011 [41]; Lavelle *et al*., 2017 [43]; Vrhovnik, 2012 [47], Kennedy *et al*., 2019 [44]; Martins *et al*., 2019 [45]). Internal consistency reliability is a measurement of the extent to which individual items of the instrument are correlated and produce consistent results of a concept or construct, through Cronbach’s alpha coefficient [28].

Three studies showed insufficient results for Cronbach’s alpha (Kennedy *et al*., 2019 [44]; Michaud, 2007 [37]; Jomori *et al*., 2017 [40]). Two of them were studies aiming to validate the cooking skills CWC questionnaire: Michaud’s evaluation tool (2007) [37] showed inadequate Cronbach’s alpha coefficients for the *Cooking Behavior scale (CB scale)*. Similar results were observed in Jomori *et al*.’s (2017) [40] study: The *Cooking Behavior (CB*) and *Cooking Attitude (CA)* scales showed low internal consistency reliability. The later authors argued that problems in the process of cross-cultural adaptations concerning translation of the original instrument into Brazilian Portuguese might have occurred. The items corresponding to these scales might not represent the constructs the authors intended to measure [28]. Thus, it is important to adjust these items for more appropriate translation into Brazilian Portuguese and to perform factor analysis [30].

Barton *et al*. (2011) [41] did not test three out of five sections of their questionnaire, under the justification that the domains within each section of the instrument assessed different constructs. Vrhovnik (2012) [47] tested only one section of her instrument to report internal consistency reliability, which the author affirmed to be reliable; however, no statistical results were shown. Therefore, we deemed these studies inadequate, according to internal consistency reliability quality criteria presented in this review.

We considered indeterminate quality of stability reported in six studies, due to insufficient sample size or inadequate time interval to perform test-retest reliability (Michaud, 2007 [37]; Warmin, 2009 [38]; Condrasky *et al*., 2011 [39]; Barton *et al*., 2011 [41]; Hartmann *et al*., 2013 [46]; Lavelle *et al*., 2017 [43]). Hartmann *et al*. (2013) [46] performed the test-retest within 1-year time interval, which may result in a measurement error to show the instrument’s stability and reproducibility [23]. Participants might improve their culinary skills during the interval between the test and the retest, especially if the elapsed time is too long. We also observed insufficient *Kappa* values (<0.7) in two studies that reported test-retest reliability. Therefore, we rated inadequate quality of this attribute.

Studies that relied exclusively on internal consistency reliability and stability analysis, without performing other psychometric measurements to validate their instruments, may not provide trustworthy results because these instruments reproduce only a consistent result in time and space from different observer (reliability), without measuring exactly what they propose (validity) [34;53]. Six studies fit into this scenario (Warmin, 2009 [38]; Condrasky *et al*., 2013 [48]; Hartmann *et al*., 2013 [46]; Barton *et al*., 2011 [41]; Kennedy *et al*., 2019 [44]; Martins *et al*., 2019 [45]). The authors only reported results for Cronbach’s alpha and test-retest reliability, and conducted inappropriate analysis for content validity, disregarding empirical evidence for experts’ agreement (or did not perform any tests for content validity). Moreover, these studies did not present any tests for construct or criterion validity.

All studies aiming to develop and validate an original instrument failed to show proper content validity: most studies relied on face validity, literature research, and experts’ judgment; however, the authors did not calculate any index to confirm experts’ agreement. Content validity based on the use of statistical methods derived from the experts’ judgment, proves itself to be essential. Otherwise, the mere fact that the experts report on the lack or excess of items representative of the construct, or that they simply determine to what extent each element corresponds to the latent phenomena, does not itself provide relevant information for the validation process [22; 28; 39, 54].

We evaluated the quality of construct validity measures of studies reporting structural validity (Michaud, 2007 [37]; Condrasky *et al*., 2011 [39]; Vrhovnik, 2012 [47]; Lavelle, *et al*., 2017 [43]; Kowalkowska, *et al*., 2018 [42]), hypothesis testing (Lavelle *et al*., 2017 [43]; Jomori *et al*., 2017 [40]) or cross-cultural validity for adapted instruments (Jomori *et al*., 2017 [40]; Kowalkowska *et al*., 2018 [42]). We observed a number of limitations, according to the quality criteria for this attribute, presented in this review.

Regarding structural validity, two studies performed principal component analysis and three studies performed exploratory factor analysis. No studies performed Confirmatory Factor Analysis (CFA). According to Gruijters, 2019 [55], exploratory factor analysis (EFA) and principal component analysis (PCA) explain correlations between items to some extent, but component analysis does a poorer job at it because it includes a portion of irrelevant variance in the analysis. If researchers have a clear idea about what a scale is supposed to be measuring, it is highly recommended studies perform Confirmatory factor analysis (CFA) to test *a priori* ideas about the latent variables researchers intend to measure [31; 30].

Only two out of five studies reporting structural validity (Lavelle *et al*., 2017 [43]; Kowalkowska *et al*., 2018 [42]) described the Kaiser-Meyer-Olkin (KMO) adequacy test with values > 0.80, which is considered very good for factor analysis appropriateness [28].

Michaud (2007) [37] performed exploratory factor analysis on insufficient sample size (minimum ratio of 5:1). Costello & Osbourne, 2005 [56] caution researchers to remember that *EFA is a “large-sample” procedure and that generalizable or replicable results are unlikely if the sample is too small*.

The cross-cultural adaptation of Michaud’s (2007) [37] instrument, reported by Jomori *et al*. (2017) [40] was adequately performed and showed satisfactory results. However, we considered inadequate quality of measure for discriminant validity between known groups, since the study showed unsupported results for significant differences between the means scores of one scale (*Self-Efficacy for Using Basic Cooking Techniques (SECT)*). This type of validity evaluates the presence of differences in the measurements obtained between the groups, not whether the measure actually measures the intended construct [57], hence, we suggest performing structural analysis to confirm construct validity of this instrument.

Vrhovnik (2012) did not provide statistical results for items factor loadings, which may imply inadequate decisions regarding retention or exclusion of an item [28].

Despite satisfactory results for convergent validity in Lavelle *et al*.’s study (2017) [43], the exploratory factor analysis (EFA) performed to validate the construct of the Cooking skills and the Food skills confidence measures showed some limitations. Four ‘Food skill’ items had higher loadings in the ‘Cooking Skills’ domain (*Buying in Season; Using leftovers to create another meal; Keeping Basics in the cupboard and Reading the best before date*), however they were retained in the ‘Food Skills’ factor. When a variable is found to have more than one significant loading, it is hard to make those factors be distinct and represent separate concepts [28]. If an instrument shows items with several cross-loadings, the items may be poorly written or the *a priori* factor structure could be flawed [58]. Moreover, two ‘Cooking Skills’ items fit into a third factor; however, they were left in the ‘Cooking Skills’ measure. One of these items consisted in ‘*Microwave food (not drinks/liquid) including heating ready-meals*’. The Brazilian Food Guide (2014) states that although microwaving may be used in meal preparation (for example microwaving a ready meal) it is not seen as a cooking skill.

Kowalkowska *et al*., 2018 [42] performed a cross cultural adaptation of Hartmann *et al*.’s cooking skills instrument (2013) [46]. However back translation was inadequately performed. Beaton (2000) [33] recommends minimum of two back-translators with the source language as their mother tongue. The main reasons are to avoid information bias and to increase the likelihood of highlighting the imperfections in the translated questionnaire.

Regarding criterion validity, little information was available in the included studies. Only one study (Michaud, 2007) presented criterion validity (predictive validity). However, we considered inadequate quality of this attribute. These findings were expected since most of the time, the criterion validity is a challenge for the researcher, because it demands a “gold standard” measure to be compared with the chosen instrument, which cannot be easily found in all knowledge areas [21; 59].

### Limitations

This review has some limitations. It is possible that some studies were missed out because they were not indexed in the databases searched, or were published for institutions, foundations, or societies. In addition, although the criteria were adapted from previous studies, the difficulty of interpreting the studies may have under- or overestimated the quality of the instruments’ psychometric properties.

## Conclusion

This review identified many studies surveying culinary skills; we considered most instruments insufficient, according to the quality of their psychometric properties. Thus, the flaws observed in these studies show that there is a need for ongoing research in the area of the psychometric properties of instruments assessing culinary skills. Moreover, our findings contribute to supporting the selection of valid and reliable instruments by healthcare professionals in clinical and Public Health settings.

Measuring culinary skills involves several separate but related domains, which integrate other constructs related to the culinary practices. Therefore, it is recommended that a more consistent and consensual definition of culinary skills as a construct be generated. Instruments should cover items and domains without overestimating one’s skills, based on his/hers ability of heating convenience food. Considering items measuring culinary skills related to the use of using basic ingredients and seasoning proves itself essential for greater understanding of barriers and facilitators related to healthy culinary practices.

## Data Availability

All data underlying the findings described fully available, without restriction.
All relevant data are within the manuscript and its Supporting Information files

## Acknowledgements

We thank the team of librarians of the School of Public Health (University of Sao Paulo) for the specialized support in electronic databases and the research group of the Department of Nutrition and Public Health of the School of Public Health (University of Sao Paulo) for proof reading the article.

## Supporting Information

**S1 Appendix**. Systematic review protocol. From: Aline Rissatto Teixeira, Daniela Bicalho, Tacio de Mendonça Lima. Evidence for the validation quality of culinary skills instruments: a systematic review. PROSPERO 2019 CRD42019130836. Available from: https://www.crd.york.ac.uk/prospero/display_record.php?ID=CRD42019130836

**S1 Table. PRISMA 2009 Checklist**. From: Moher D, Liberati A, Tetzlaff J, Altman DG. The PRISMA Group (2009). Preferred Reporting Items for Systematic Reviews and Meta-Analyses: The PRISMA Statement. PLoS Med 6(7): e1000097. doi:10.1371/journal.pmed1000097

**S2 Appendix**. Search strategy until January 12, 2021.

**S2 Table**. List of excluded studies.

