## Supplementary material for "Systematic review of instruments for assessing culinary skills in adults: What is the quality of their psychometric properties?": S2 Appendix

**S2 Appendix.** Search strategy until January 12, 2021.

**A. Scopus (n = 359)**

TITLE-ABS-KEY(Cooking OR Cookery OR "culinary skill*" OR "cooking skill*" OR "cooking abilit*") AND TITLE-ABS-KEY("Survey Methods" OR "Survey Method" OR "Survey Methodology" OR "Community Surveys" OR "Community Survey" OR "Repeated Rounds of Survey" OR Surveys OR Survey OR "Questionnaire Design" OR "Questionnaire Designs" OR "Baseline Survey" OR "Baseline Surveys" OR Respondents OR Respondent OR "Randomized Response Technique" OR "Randomized Response Techniques" OR Questionnaires OR Questionnaire OR Nonrespondents OR Nonrespondent OR Scale* OR Instrument*) AND TITLE-ABS-KEY("Validation Study" OR "Validation Studies" OR "Reproducibility of Results" OR "Reproducibility of Findings" OR "Reproducibility Of Result" OR "Reproducibility of Finding" OR "Finding Reproducibilities" OR "Finding Reproducibility" OR "Reliability of Results" OR "Reliability of Result" OR "Result Reliabilities" OR "Result Reliability" OR Reliability OR Validity OR "Validity of Results" OR "Validity of Result" OR "Result Validities" OR "Result Validity" OR "Face Validity" OR "Reliability and Validity" OR "Validity and Reliability" OR "Test-Retest Reliability" OR "Test Retest Reliability" OR "Factor Analysis" OR "Statistical Factor Analyses" OR "Statistical Factor Analysis" OR "Factor Analyses" OR "Consensus" OR "Consensus Development")

**B. LILACS (n = 7)**

#1 MH:Cooking OR (Cooking) OR (Cookery) OR (culinary skill$) OR (cooking skill$) OR (cooking abilit$) OR J01.576.423.200.200$

#2 MH:Surveys and Questionnaires OR (Surveys and Questionnaires) OR (Questionnaires and Surveys) OR (Survey Methods) OR (Methods, Survey) OR (Survey Method) OR (Methodology, Survey) OR (Survey Methodology) OR (Community Surveys) OR (Community Survey) OR (Survey, Community) OR (Surveys, Community) OR (Repeated Rounds of Survey) OR (Surveys) OR (Survey) OR (Questionnaire Design) OR (Design, Questionnaire) OR (Designs, Questionnaire) OR (Questionnaire Designs) OR (Baseline Survey) OR (Baseline Surveys) OR (Survey, Baseline) OR (Surveys, Baseline) OR (Respondents) OR (Respondent) OR (Randomized Response Technique) OR (Randomized Response Techniques) OR (Response Technique, Randomized) OR (Response Techniques, Randomized) OR (Techniques, Randomized Response) OR (Questionnaires) OR (Questionnaire) OR (Nonrespondents) OR (Nonrespondent) OR (Scale$) OR (Instrument$) OR E05.318.308.980$ OR N05.715.360.300.800$ OR N06.850.520.308.980$

#1 AND #2

**C. PubMed (624)**

#1 "Cooking"[Mesh] OR (Cooking) OR (Cookery) OR (culinary skill*) OR (cooking skill*) OR (cooking ability*)

#2 "Surveys and Questionnaires"[Mesh] OR (Surveys and Questionnaires) OR (Questionnaires and Surveys) OR (Survey Methods) OR (Methods, Survey) OR (Survey Method) OR (Methodology, Survey) OR (Survey Methodology) OR (Community Surveys) OR (Community Survey) OR (Survey, Community) OR (Surveys, Community) OR (Repeated Rounds of Survey) OR (Surveys) OR (Survey) OR (Questionnaire Design) OR (Design, Questionnaire) OR (Designs, Questionnaire) OR (Questionnaire Designs) OR (Baseline Survey) OR (Baseline Surveys) OR (Survey, Baseline) OR (Surveys, Baseline) OR (Respondents) OR (Respondent) OR (Randomized Response Technique) OR (Randomized Response Techniques) OR (Response Technique, Randomized) OR (Response Techniques, Randomized) OR (Techniques, Randomized Response) OR (Questionnaires) OR (Questionnaire) OR (Nonrespondents) OR (Nonrespondent) OR (Scale*) OR (Instrument*)

#3 "Validation Study" [Publication Type] OR (Validation Study) OR "Validation Studies as Topic"[Mesh] OR (Validation Studies as Topic) OR "Reproducibility of Results"[Mesh] OR (Reproducibility of Results) OR (Reproducibility of Findings) OR (Reproducibility Of Result) OR (Of Result, Reproducibility) OR (Of Results, Reproducibility) OR (Result, Reproducibility Of) OR (Results, Reproducibility Of) OR (Reproducibility of Finding) OR (Finding Reproducibilities) OR (Finding Reproducibility) OR (Reliability of Results) OR (Reliability of Result) OR (Result

Reliabilities) OR (Result Reliability) OR (Reliability (Epidemiology)) OR (Validity (Epidemiology)) OR (Validity of Results) OR (Validity of Result) OR (Result Validities) OR (Result Validity) OR (Face Validity) OR (Validity, Face) OR (Reliability and Validity) OR (Validity and Reliability) OR (Test-Retest Reliability) OR (Reliabilities, Test-Retest) OR (Reliability, Test-Retest) OR (Test Retest Reliability) OR "Factor Analysis, Statistical"[Mesh] OR (Factor Analysis, Statistical) OR (Analysis, Statistical Factor) OR (Analyses, Statistical Factor) OR (Factor Analyses, Statistical) OR (Statistical Factor Analyses) OR (Statistical Factor Analysis) OR (Analysis, Factor) OR (Analyses, Factor) OR (Factor Analyses) OR (Factor Analysis) OR "Consensus"[Mesh] OR (Consensus) OR (Consensus Development) OR (Development, Consensus)

#1 AND #2 AND #3

**Web of Science (487)**

#1 TS=(Cooking OR Cookery OR "culinary skill*" OR "cooking skill*" OR "cooking abilit*")

#2 TS=("Survey Methods" OR "Survey Method" OR "Survey Methodology" OR "Community Surveys" OR "Community Survey" OR "Repeated Rounds of Survey" OR Surveys OR Survey OR "Questionnaire Design" OR "Questionnaire Designs" OR "Baseline Survey" OR "Baseline Surveys" OR Respondents OR Respondent OR "Randomized Response Technique" OR "Randomized Response Techniques" OR

Questionnaires OR Questionnaire OR Nonrespondents OR Nonrespondent OR Scale* OR Instrument*)

#3 TS=(“Validation Study” OR “Validation Studies” OR “Reproducibility of Results” OR “Reproducibility of Findings” OR “Reproducibility Of Result” OR “Reproducibility of Finding” OR “Finding Reproducibilities” OR “Finding Reproducibility” OR “Reliability of Results” OR “Reliability of Result” OR “Result Reliabilities” OR “Result Reliability” OR Reliability OR Validity OR “Validity of Results” OR “Validity of Result” OR “Result Validities” OR “Result Validity” OR “Face Validity” OR “Reliability and Validity” OR “Validity and Reliability” OR “Test-Retest Reliability” OR “Test Retest Reliability” OR "Factor Analysis" OR “Statistical Factor Analyses” OR “Statistical Factor Analysis” OR “Factor Analyses” OR "Consensus" OR “Consensus Development”)

#1 AND #2 AND #3

**D. Google Scholar (n = 106)**

"culinary skills" OR "cooking skills" OR "cooking abilities" AND scale OR instrument* OR survey OR questionnaire* AND "Validation Studies"

Filters:

- No citations and patents
