## Supplementary material for "Systematic review of instruments for assessing culinary skills in adults: What is the quality of their psychometric properties?": S2 Table

**S2 Table.** List of excluded studies.

| Reason for exclusion | Author, Year | Title | Reference |
|---|---|---|---|
| Wrong outcome | Bech Larsen & Tsalis, 2018 | Impact of cooking competence on satisfaction with food-related life: Construction and validation of cumulative experience & knowledge scales | Food Quality and Preference. 68: 191–197. doi: 10.1016/j.foodqual.2018.02.006 |
| Wrong outcome | Bongoni, et.al , 2015 | Evaluation of research methods to study domestic food preparation | British Food Journal. 117(1):7-21 doi: 10.1108/BFJ-09-2013-0273 |
| Wrong outcome | Garaham et. al, 2013 | Perceived Social Ecological Factors Associated with Fruit and Vegetable Purchasing, Preparation, and Consumption among Young Adults | Journal of the American Academy of Nutrition and Dietetics. 113(10) doi:10.1016/j.jand.2013.06.348 |
| Wrong outcome | Ko, 2010 | To Evaluate the Professional Culinary Competence of Hospitality Students | Journal of Culinary Science & Technology. 8(2):136-146 doi: 10.1080/15428052.2010.511101 |
| Wrong outcome | Lane, et. al, 2017 | Development of the Cooking and Food Provisioning Action Scale (CAFPAS): A new measurement tool for individual cooking practice | Food Quality and Preference. 62(2) doi: 10.1016/j.foodqual.2017.06.022 |
| Wrong outcome | Pinard et. al, 2018 | Development and Testing of a Revised Cooking Matters for Adults Survey | American Journal of Health Behavior. 39(6):866-873 doi: 10.5993/AJHB.39.6.14 |
| Wrong outcome | Poncet et. al, 2015 | Reliability of the Cooking Task in adults with acquired brain injury | Neuropsychological Rehabilitation. 25(2):298-317. doi: 10.1080/09602011.2014.971819. |
