## Supplementary material for "Systematic review of instruments for assessing culinary skills in adults: What is the quality of their psychometric properties?": S1 Appendix

### Citation

Aline Rissatto Teixeira, Daniela Bicalho, Tacio de Mendonça Lima. Evidence for the validation quality of culinary skills instruments: a systematic review. PROSPERO 2019 CRD42019130836 Available from: https://www.crd.york.ac.uk/prospero/display_record.php?ID=CRD42019130836

### Review question

What is the scientific evidence for the quality of validated instruments for evaluating culinary skills?

### Searches

We will seach the following electronic bibliographic databases: Scopus, LILACS, PubMed, Web of Science, SIBiUSP Integrated Search Portal and the grey literature (through DOAJ and Google Scholar).

In addition, the secondary references of the included articles will also be analyzed for further material.

The search strategy will include only terms relating to culinary / cooking / food skills / confidence / knowledge OR cooking abilities OR food literacy OR food agency OR food autonomy AND validation studies OR validation OR pshychometrics OR scale OR instrument OR survey.

There will be no publication period restrictions.

Studies published in English, Portuguese or Spanish will be eligible for inclusion.

Additional search strategy information can be found in the attached PDF document (link provided below).

### Types of study to be included

Inclusion:

We will include methodological studies with a psychometric approach if:
1) They present an original instrument that evaluates of culinary skills in adults (20 years of age or older and under 60, according to Brazilian Ministry of Health);

2) They present the development of such instrument based on bibliographic references combined or not with group discussions;

3) They describe the process of validation and reliability of the instrument;

4) They are published in English, Portuguese or Spanish.

Exclusion:

Instruments tested in university students, children and adolescents or that do not present the instrument will be excluded.

### Condition or domain being studied

Culinary skills; food literacy; food agencies.

Culinary skills have been valued in public policies and national health instruments, such as the Brazilian Food Guide, and seem to be important for the individual, who must feel empowered and confident to make healthy and autonomous food choices.
Although the number of studies about this phenomenon is increasing in the last years, it is necessary to describe the evaluation of the culinary abilities in a precise way, starting from the use of validated instruments or even discussing the necessary criteria to evidence its validation.

Criteria to evaluate psychometric quality of instruments address properties such as reliability (including internal consistency), and validity (including content, construct and criteria).

### Participants/population

Adults (20 years of age or older and under 60, according to Brazilian Ministry of Health).

Articles for the validation of instruments tested in university students, children and adolescents or that do not present the instrument will be excluded.

### Intervention(s), exposure(s)

Validation quality of culinary skills assessment instruments.

### Comparator(s)/control

Not applicable.

### Main outcome(s)

A psychometric qualification of two instruments proposed will be carried out using a system of classification adapted from Terwee et al, and Hair Jr. et al.

The questions used for this classification address the following properties:

a) Reliability, including internal consistency;

b) Validity, including reference to the content, construction and criteria.

#### * Measures of effect

Not applicable.

### Additional outcome(s)

None.

#### * Measures of effect

Not applicable.

### Data extraction (selection and coding)

After defining the descriptors and searching for articles in the databases, the results will be screened for relevance.

The titles and abstracts of the retrieved studies will be independently screened and selected by two authors. Studies not meeting the inclusion criteria will be eliminated from the review. For this process, a preformatted Microsoft Excel worksheet will be used. The selected articles will be grouped by database and repeated article titles eliminated. Then, a consensus will be reached by classifying the selected articles in: Yes (enter in review), No (do not enter in review) and not sure (doubtful).

Any disagreements between the researchers will be resolved by a third reviewer.

After consensus has been achieved, studies classified as "No" will be excluded.

The full texts of the included articles will then be obtained and read. If they are not available in the databases, the authors will be contacted by e-mail or other tools, such as www.researchgate.com. The selected studies will be checked to certify that they answer the guiding question of the review, that the eligibility criteria have been met. Any not fulfilling these criteria will be rejected.

Any disagreements between the researchers will be resolved by a third reviewer.

Secondary references will also be analyzed.

Data extraction will then be conducted on the studies selected for inclusion independently by two reviewers, again using a preformatted Microsoft Excel worksheet. Any disagreements between the researchers will also, again, be resolved by a third reviewer. For each included study, the extracted information will consist of: the country, participants, setting, sample size, format of instrument, target public, number of items of the instrument, instrument development, the instrument domains, and the instrument psychometric properties.

### Risk of bias (quality) assessment

The characteristics of reliability and validity are very important in the development or adaptation of research-measuring instruments, considering that studies may present consistent results because they may be flawed or exhibit missing data. Therefore, the risk of bias will be assessed according to the rating system adapted from Terwee et al (2007) and Hair Jr, et al (2009).

The criteria for this rating system address the following properties:

a) Reliability, including internal consistency (Cronbach's alpha between 0.70 and 0.95);

b) Validity, including: content (description provided about the aims of the instrument, target population, concepts that are being measured, item selection and about the investigators or experts involved for the items selection), construct (Factor analyses performed on adequate sample size (minimum ration of 5:1and > 100), and Bartlett's sphericity test (p < 0.005) or KMO adequacy test (>0.7) or factors explaining > 60% of the variance or RMSEA < 0, 07 or GFI and AGFI > 0.95 or SRMR <0, 08 or CFI > 0.95), and criterion validity (with convincing arguments that gold standard for used comparison with the developed instrument is ‘‘gold’’ and correlation with gold standard is >0.70).

Two independent authors will apply this rating system. Any divergences between researchers will be resolved by a third reviewer.

### Strategy for data synthesis

We will provide a narrative synthesis of the findings from the included studies, structured around the type of methodology applied, and the target population characteristics.

We will provide summaries and comparison of each study outcomes with the recommendations established by authors referenced in psychometric literature.

Selected studies will be categorized by the country of origin, the language in which the study has been published, the number and type of participants, the type of developed instrument (i.e. scale, questionnaire), the instrument domains, and the applied methods for psychometric validation.

The analysis of the psychometric quality of the instruments will address the following properties: reliability, including internal consistency and stability;and validity, including , content, construct, and criterion validity.

Each measurement property will be reported by positive, (+), intermediate (?), negative (-), or no information available (0) and the data will be organized in a preformatted Microsoft Excel worksheet, and reported in a table.

### Analysis of subgroups or subsets

Selected studies will be categorized by the country of origin, the language in which the study has been published, the number and type of participants, the type of developed instrument (i.e. scale, questionnaire), the instrument domains, and the applied methods for psychometric valitation.

The analysis of the psychometric quality of the instruments will address the following properties: reliability, including internal consistency and validity, including, content, construct, and criterion validity.

### Contact details for further information

Aline Rissatto Teixeira


### Organisational affiliation of the review

Universidade de São Paulo
https://www.fsp.usp.br

### Review team members and their organisational affiliations

Ms Aline Rissatto Teixeira. Universidade de São Paulo
Ms Daniela Bicalho. Universidade de São Paulo
Mr Tacio de Mendonça Lima. Universidade de São Paulo

### Collaborators

Mrs Betzabeth Slater Villar. Universidade de São Paulo

### Type and method of review

Methodology, Systematic review

### Anticipated or actual start date

01 May 2019

### Anticipated completion date

21 December 2019

### Funding sources/sponsors

None

### Conflicts of interest

### Language

English, Portuguese-Brazil, Portuguese-Local, Spanish

### Country

Brazil

### Stage of review

Review Ongoing

### Subject index terms status

Subject indexing assigned by CRD

### Subject index terms

Cooking; Food; Healthy Diet; Humans; Psychometrics; Reproducibility of Results; Research Design

### Date of registration in PROSPERO

10 June 2019

### Date of first submission

23 April 2019

### Stage of review at time of this submission

The review has not started

| Stage | Started | Completed |
|---|---|---|
| Preliminary searches | No | No |
| Piloting of the study selection process | No | No |
| Formal screening of search results against eligibility criteria | No | No |
| Data extraction | No | No |
| Risk of bias (quality) assessment | No | No |
| Data analysis | No | No |

*The record owner confirms that the information they have supplied for this submission is accurate and complete and they understand that deliberate provision of inaccurate information or omission of data may be construed as scientific misconduct.*

*The record owner confirms that they will update the status of the review when it is completed and will add publication details in due course.*

### Versions

10 June 2019

PROSPERO
This information has been provided by the named contact for this review. CRD has accepted this information in good faith and registered the review in PROSPERO. The registrant confirms that the information supplied for this submission is accurate and complete. CRD bears no responsibility or liability for the content of this registration record, any associated files or external websites.
